# Glucose and lipid profiles in men with non-obstructive azoospermia

**DOI:** 10.1101/2022.03.14.22272336

**Authors:** Ponco Birowo, Dimas Tri Prasetyo, Dwi Ari Pujianto, Widi Atmoko, Nur Rasyid

**Affiliations:** Department of Urology, Faculty of Medicine Universitas Indonesia - Dr. Cipto Mangunkusumo Hospital, Jakarta; Department of Medical Biology, Faculty of Medicine Universitas Indonesia, Jakarta

**Keywords:** azoospermia, NOA, OA, serum testosterone, metabolic profile

## Abstract

The impact of a low testosterone level among men with non-obstructive azoospermia with various testicular histopathological patterns on the regulation of glucose and lipid metabolism is less well known than among the general population. The aim of this retrospective study was to examine the association between testicular histopathology and components of the metabolic profile among men with non-obstructive azoospermia. Participants were divided into two groups: men with non-obstructive azoospermia and men with obstructive azoospermia. Testicular biopsies were performed among those with non-obstructive azoospermia. We included 115 patients in this study: 83 (72.2%) had non-obstructive azoospermia and 32 (27.8%) had obstructive azoospermia. The plasma glucose concentration, glycated hemoglobin level, and lipid profile were similar between patients with non-obstructive azoospermia and those with obstructive azoospermia. Upon subgroup analysis of patients with non-obstructive azoospermia, those with Sertoli-cell-only syndrome had the lowest testosterone (431 ± 238 ng/dL; *P*=0.039) and highest follicle-stimulating hormone (23.4 ± 18.2 mIU/mL; *P=*0.002) concentrations. They also had the highest triglyceride concentration (163 ± 114 mg/dL; *P*=0.001). Interestingly, patients with Sertoli-cell-only syndrome had a lower fasting plasma glucose concentration (92 ± 11 mg/dL; *P*<0.001) and glycated hemoglobin level (5.9 ± 0.8%; *P*=0.022) than those with histopathological patterns of maturation arrest and hypospermatogenesis. In conclusion, differences in glucose and lipid metabolism are evident between men with non-obstructive azoospermia with different spermatogenesis patterns.

## Introduction

The association between testosterone and the development of type-2 diabetes mellitus (DM), obesity, and dyslipidemia has been demonstrated [1–6]. Testosterone deficiency is reportedly related to metabolic syndrome [7]. The exact mechanism by which testosterone may affect the metabolic profile is yet to be established.

Testosterone is thought to have protective effects against the pathogenesis of metabolic syndrome and its components by increasing skeletal muscle tissue and decreasing abdominal obesity and the levels of nonesterified fatty acids in the body [8]. Testosterone reportedly has a favorable effect on vascular reactivity, inflammation, cytokine production, and the expression of adhesion molecules, as well as on the serum lipid concentration and hemostatic factors [9].

Despite the fact that there have been numerous studies in which the association between testosterone and the metabolic profile has been investigated, its effect on the regulation of glucose and lipid metabolism in infertile men with non-obstructive azoospermia (NOA), particularly in relation to testicular histopathology, is less well known. Thus, the aim of this study was to examine the effect of testosterone level on the plasma glucose and lipid profiles of men with NOA, as well as its impact based on testicular histopathology patterns. We revealed that the presence of testicular damage in NOA is associated with lower serum testosterone and higher levels of follicle-stimulating hormone (FSH), as well as lower fasting plasma glucose and higher triglyceride levels than the absence of such damage.

## Material and methods

### Patient selection

In this retrospective study, we included all men with azoospermia seeking infertility treatment at the Urology Clinics of Dr. Cipto Mangunkusumo Hospital and Bunda Hospital (Jakarta, Indonesia) from 2016 to 2020. Azoospermia is defined as the absence of spermatozoa in the seminal fluid after centrifugation. The diagnosis was made only when no spermatozoa was found in two consecutive semen analyses, each after 2-7 days of sexual abstinence. Patients who were recruited into this study did not have any prior history of type-2 DM or dyslipidemia and did not take any medication that may affect glucose or lipid metabolism in the 3 months before recruitment. Patients were excluded if had any prior history of type-2 DM, dyslipidemia, and taking any medication that may affect glucose or lipid metabolism in the 3 months before recruitment. Type-2 DM is defined as 1) having a fasting plasma glucose concentration of ≥126 mg/dL, or 2) 2-hour plasma glucose ≥200 mg/dL (11.1 mmol/L) during oral glucose tolerance test, or 3) A1C ≥6.5% (48 mmol/mol), or 4) experiencing classical symptoms of hyperglycemia or hyperglycemic crisis (polyuria or polydipsia) and having a random plasma glucose concentration ≥200 mg/dL [10]. Dyslipidemia is defined as a history of exhibiting either a total cholesterol concentration ≥200 mg/dL, a high-density lipoprotein (HDL) cholesterol concentration <40 mg/dL for men and <50 mg/dL for women, or a triglyceride concentration >150 mg/dL [11]. There were two study groups in this study: patients with NOA and those with obstructive azoospermia (OA). The NOA/OA diagnosis was based on the patient history, clinical examination, and reproductive hormone (follicle-stimulating hormone [FSH], luteinizing hormone, and testosterone) assessment, and confirmed in a testicular histopathology specimen taken during sperm retrieval procedures. This study was approved by the Ethics Committee of the Faculty of Medicine, Universitas Indonesia and complies with the Code of Ethics of the World Medical Association (Declaration of Helsinki 1964, revised 20033 and Declaration of Tokyo 1975, revised 20064). Informed consent from participants were not required due to retrospective data collection.

### Study design

We conducted hormonal and metabolic evaluations of all participants, which included concentrations of serum testosterone, serum FSH, fasting plasma glucose, glycated hemoglobin (HbA1c), total cholesterol, low-density lipoprotein (LDL), high-density lipoprotein (HDL), and triglyceride. Participants were asked to fast for 10-12 h before their blood was drawn. Serum testosterone was examined by using an electrochemiluminescence immunoassay (Roche, Basel, Switzerland). Serum FSH was examined by using immunochemiluminescence (Siemens Healthineers, Erlangen, Germany). Fasting plasma glucose was measured by using the hexokinase method (Diatron Promedika, Jakarta, Indonesia). HbA1c was examined by using an enzymatic assay (Abbott Architect, Illinois, USA). Total cholesterol was measured by using the cholesterol oxidase-peroxidase aminoantipyrine technique, LDL and HDL were examined by using homogeneous assays, and triglyceride was measured by using glycerol peroxidase phosphate method (all reagents for lipid profiling were from Diatron Promedika). The serum testosterone concentration was presented as nanograms/deciliter (ng/dL), the FSH concentration was presented as milli-international units per milliliter (mIU/mL), while the fasting plasma glucose, total cholesterol, LDL cholesterol, HDL cholesterol, and triglyceride concentrations were presented as milligrams/deciliter (mg/dL). HbA1c was presented as a percentage (%).

### Testicular biopsy

We also performed surgical sperm retrieval by using testicular sperm extraction (TESE) for patients with NOA as a part of their infertility treatment. TESE was performed by making a small incision on the avascular plane of the testis and collecting a small portion of the testicular tissue for examination for the presence of motile spermatozoa under an electric light microscope.

We also performed a second testicular biopsy for histological examination of the testicular phenotype (spermatogenesis pattern). Samples were added into a Bouin’s solution and sent to the laboratory for examination. In the laboratory, several sections of the samples were prepared and stained in paraffin blocks. The section with the highest concentration of seminiferous tubules was examined by the same laboratory analyst for all samples under a light microscope. Testicular histopathological patterns were divided into three categories (Fig 1): (i) Sertoli cell only (SCO; absence of germ cells in the seminiferous tubules), (ii) maturation arrest (incomplete spermatogenesis), and (iii) hypospermatogenesis (a reduced number of normal spermatogenic cells). All sperm retrieval and testicular biopsy procedures were performed by the same surgeon, while all testicular histopathological examinations in this study were performed in the same laboratory by the same examiner.

**Fig 1.** Classification of spermatogenesis based on testicular histopathological patterns. A. Sertoli cell only (#). B. Maturation arrest with spermatocyte cells ($). C. Hypospermatogenesis, which is marked by a decreased number of normal spermatozoa (^). Courtesy of the Department of Biology, Faculty of Medicine, Universitas Indonesia, Jakarta.

### Statistical analysis

All continuous data were presented as mean and standard deviation value as it met the normality distribution assumption by using Shapiro-Wilk test. No outlier was removed from the analysis. We compared the results of the hormonal and metabolic evaluations between patients with NOA and those with OA by using the two-tailed independent t-test. We performed subgroup analysis within the NOA group based on the testicular histopathological patterns by using one-way analysis of variance and post-hoc Bonferroni test. Minimum sample size is 30 for each group based on hypothesis testing for two mean formula (power 80%, alpha 5%) [12]. All statistical analyses were performed by using IBM SPSS Statistics for Windows version 20.0 (IBM Corp., Armonk, NY, USA) [13], with *P*<0.05 considered statistically significant.

## Results

We included 115 patients in this study, of which 83 (72.2%) were in the NOA group and 32 (27.8%) were in the OA group. The age range of patients was 26-72 years and there were no significant differences in age or body mass index between the two study groups (Table 1).

**Table 1.** Comparisons of hormonal and metabolic parameters between patients with NOA and those with OA.

| Parameter | NOA (n = 83) | OA (n = 32) | *P* value |
| --- | --- | --- | --- |
| Age (years) | 36 ± 7 | 38 ± 9 | 0.308 / *t* = 1.024 |
| BMI (kg/m<sup>2</sup>) | 26.7 ± 5.4 | 27.7 ± 5.5 | 0.632 / *t* = 0.483 |
| **Serum testosterone (ng/dL)** | **442 ± 181** | **511 ± 126** | **0.049 / *t* = 1.970** |
| **FSH (mIU/mL)** | **16.3 ± 13.8** | **4.3 ± 1.1** | **<0.001 / *t* = -4.879** |
| Fasting plasma glucose (mg/dL) | 98 ± 32 | 90 ± 10 | 0.143 / *t* = -1.476 |
| HbA1c (%) | 6.1 ± 1.4 | 5.4 ± 0.4 | 0.132 / *t* = -1.534 |
| Total cholesterol (mg/dL) | 203 ± 34 | 197 ± 53 | 0.462 / *t* = -0.738 |
| LDL (mg/dL) | 143 ± 38 | 159 ± 44 | 0.240 / *t* = 1.188 |
| HDL (mg/dL) | 42.8 ± 9.1 | 43.9 ± 8.5 | 0.574 / *t* = 0.564 |
| Triglyceride (mg/dL) | 145 ± 81 | 124 ± 44 | 0.159 / *t* = -1.417 |

*P* values were calculated with the two-tailed independent t-test and degree of freedom 113 All results are presented
as the mean ± standard deviation.
NOA = non-obstructive azoospermia; OA = obstructive azoospermia; BMI = body mass index; FSH = follicle-stimulating hormone; HbA1c = glycated hemoglobin; HDL = high-density lipoprotein; LDL = low-density lipoprotein.

Patients with NOA had significantly lower serum testosterone and higher FSH concentrations compared to those with OA (Table 1). Furthermore, no significant differences were observed in glucose and lipid profiles between the two study groups. Among patients with NOA, patients with SCO testicular histopathology had the lowest serum testosterone (431 ± 238 ng/dL; *P*=0.039) and highest FSH (23.4 ± 18.2 mIU/mL; *P*=0.002) concentrations (Table 2). Patients with SCO had a lower total cholesterol than those with maturation arrest and hypospermatogenesis (185 ± 29 mg/dL, 208 ± 35 mg/dL, and 215 ± 31 mg/dL, respectively; *P*=0.008). A comparison of LDL levels among those groups yielded similar results (122 ± 25, 160 ± 18, and 146 ± 8, respectively; *P*=0.012).

**Table 2.**
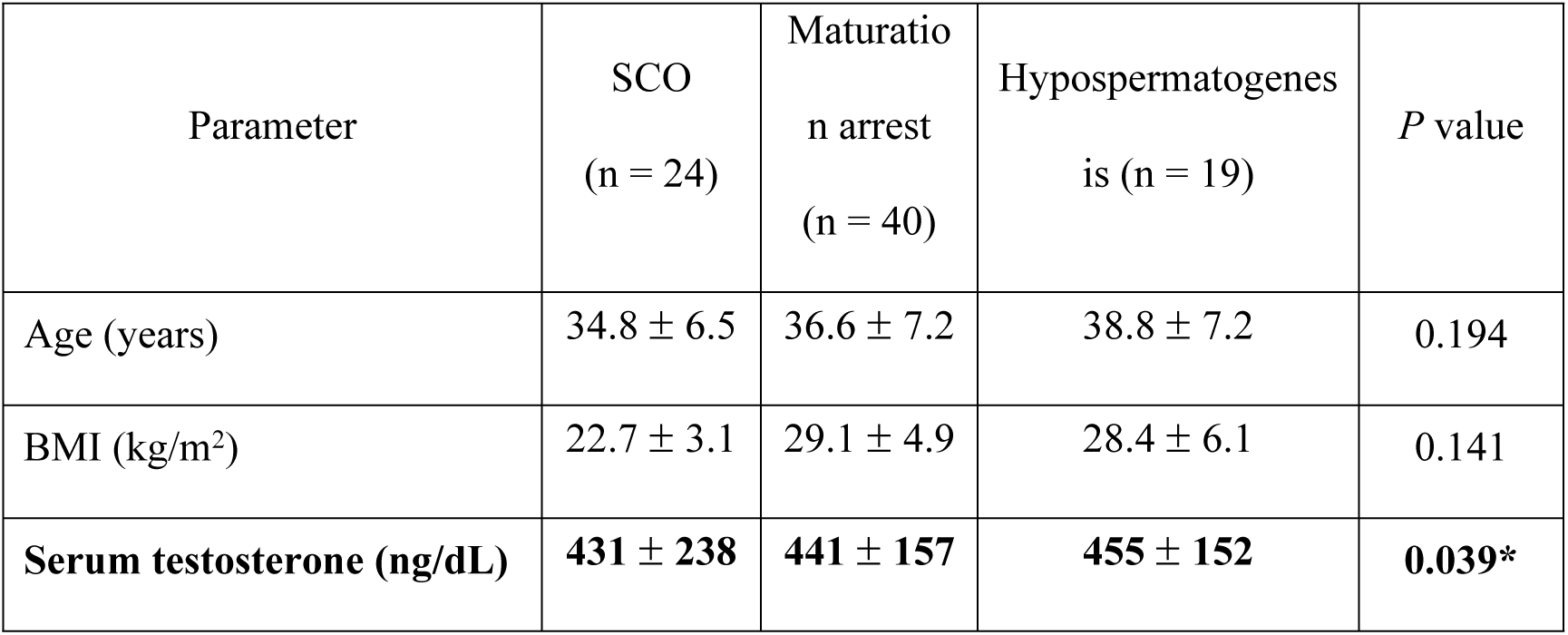
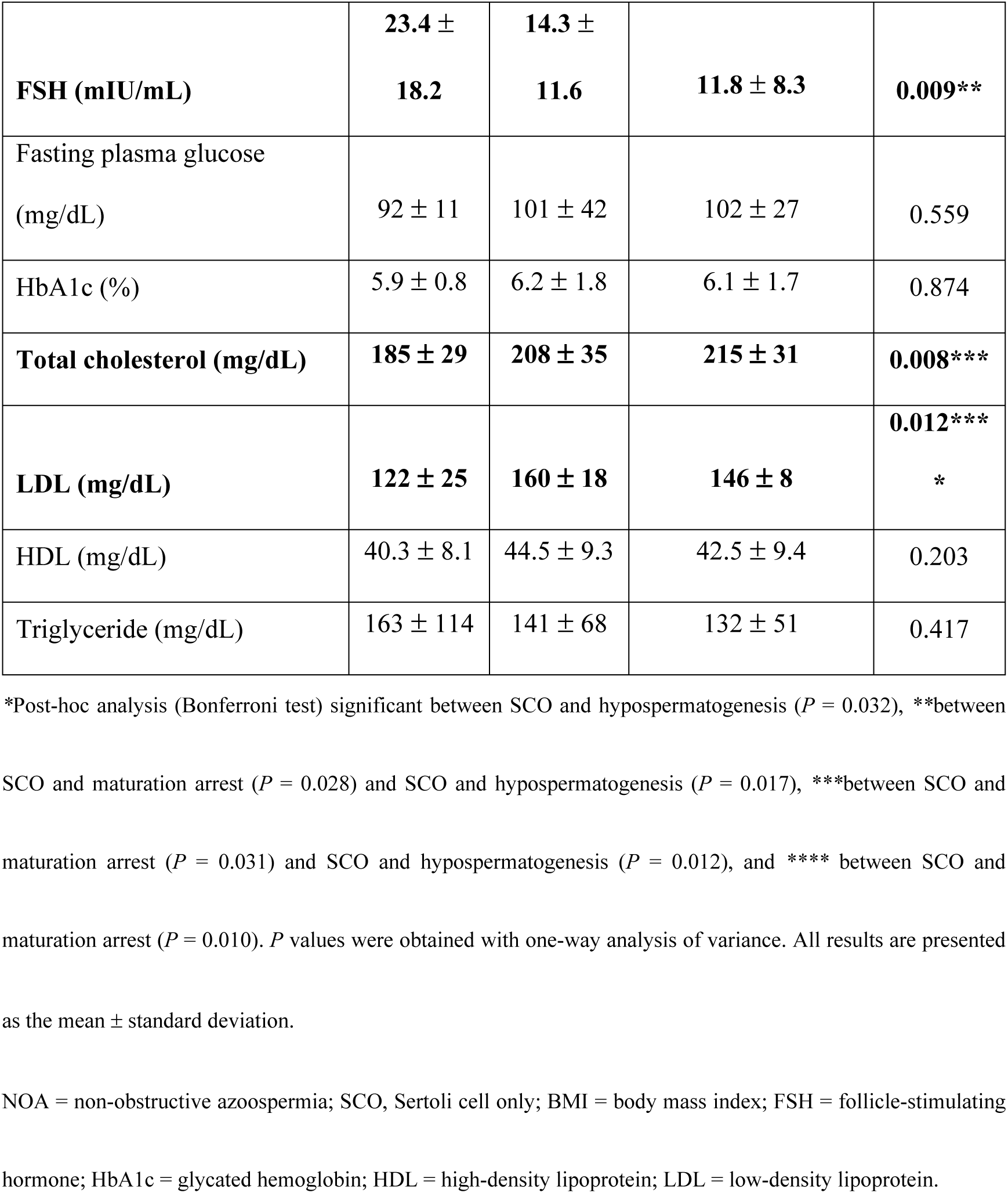
Comparisons of hormonal and metabolic parameters among patients with NOA based on testicular histopathological patterns.

## Discussion

We are aware of only a few studies in which the association between male infertility and glucose and lipid metabolism was investigated. Our results revealed that NOA was more common than OA among patients with azoospermia seeking fertility treatment at our center. This is consistent with previous studies, in which a higher incidence of NOA than OA was reported among men with azoospermia [14,15]. FSH and testosterone evaluation are important aspects of the assessment of patients with azoospermia, to, for example, differentiate between NOA and OA. Typically, men with NOA have higher FSH and lower testosterone concentrations and a more abnormal pattern of testicular histopathology than those with OA [16]. Thus, the high FSH and low testosterone values in men with NOA in our study were expected and consistent with the literature. The etiologies of and risk factors for NOA include Klinefelter’s syndrome, cryptorchidism, Y-chromosome microdeletion, and Kallmann syndrome [17–19]. The causes of NOA were not investigated in this study.

Although not statistically significant, patients with NOA had a higher triglyceride level than those with OA (145 ± 81 mg/dL vs. 124 ± 44 mg/dl; *P*=0.159). This parameter, however, exhibited statistically significant differences among patients with NOA with different patterns of spermatogenesis upon testicular histopathology. A study by Bobjer et al. [20] on patients with hypogonadism and NOA revealed that the testosterone and triglyceride concentrations were positively correlated (*P*=0.03). Hypertriglyceridemia and a low HDL concentration was also more commonly detected in men with azoospermia and oligospermia than in normal controls in a study by Mendoza et al [21]. Calonge et al. [22] also reported a correlation between triglyceride concentration and infertility, where a higher triglyceride concentration was observed in patients who were infertile compared to controls (164 vs. 91 mg/dL; *P*<0.01). None of these studies, however, demonstrated a causal relationship between dyslipidemia and azoospermia.

Despite the limited data available on the association between male infertility, particularly NOA, and glucose metabolism, many studies have demonstrated a relationship between plasma glucose concentration/development of type-2 DM and hypogonadism. Laaksonen et al. [23], who conducted a cohort study on 702 middle-aged Finnish men without a history of type-2 DM or metabolic syndrome, discovered that men with testosterone, free testosterone, and sex hormone-binding globulin concentration in the lower quartile had odds ratios ranging from 1.57 (95% confidence interval [CI]=0.98-2.50) to 2.89 (95% CI=1.93-4.33) for the development of metabolic syndrome after an 11-year follow-up, even after adjusting for age, smoking status, alcohol intake, socioeconomic status, and previous cardiovascular disease. Pitteloud et al. [22], investigating 60 men (age range: 39-65 years) without a history of testicular disorder, discovered that testosterone concentration was correlated with insulin sensitivity. Patients with a low serum testosterone concentration had a threefold higher prevalence of metabolic syndrome compared to patients with normal serum testosterone level (90% vs. 29%). Blouin et al. revealed that the effects of declining dehydroepiandrosterone sulfate, an endogenous androgen, on the metabolic profile were age dependent, unlike those of serum testosterone [24].

There are many other studies in which the association between testosterone and the metabolic profile was examined. However, most of them were focused on the effects of type-2 DM on the testosterone level. Corona et al. [25] conducted a meta-analysis on the association between testosterone and type-2 DM, discovering that patients with type-2 DM exhibited statistically significantly lower concentrations of plasma testosterone than non-diabetic individuals. The same group also revealed that both type-2 DM and metabolic syndrome were independent predictors of a low testosterone concentration [26].

In this study, patients with SCO had the lowest testosterone and highest FSH concentrations. This was in accordance with a previous study by Yassin et al. [27], wherein lower testosterone (1.7 ± 1.3 vs. 5.0 ± 2.2 ng/mL; *P*<0.001) and higher FSH (20.8 ± 14.8 vs 7.7 ± 3.9 mIU/mL; *P*<0.001) concentrations were found in patients with SCO syndrome (SCOS) than in fertile men. Another study of 134 patients with SCOS also revealed low testosterone (4.4 ± 1.6 ng/mL) and high FSH (18.9 ± 9.0 mIU/mL) concentrations [28]. Interestingly, in our study, patients with SCOS had higher total cholesterol and LDL concentration than patients with maturation arrest and those with hypospermatogenesis. We are not aware of an existing report to this effect. On the other hand, the triglyceride concentration was higher in patients with the poorest histopathologic pattern (SCO), although statistically not significant. This may indicate that triglyceride metabolism is related to the process of spermatogenesis. Our results regarding glucose and lipid metabolism among patients with NOA with different patterns of testicular histopathology, however, may not be generalizable, as the number of patients was relatively modest. Thus, further studies on the association between different levels of testicular damage and the metabolic profile are warranted, with larger study samples.

In conclusion, the presence of testicular damage in NOA is associated with lower serum testosterone and higher FSH concentrations, as well as with lower total cholesterol and LDL concentrations. Further studies are needed to better explain these phenomena, incorporating larger samples and follow-ups. All the components of metabolic syndrome, i.e., hypertension, hyperglycemia or insulin resistance, large waist circumference, and hypercholesterolemia or hypertriglyceridemia, must also be incorporated in future studies, so that the association between male infertility and the development of metabolic syndrome can be investigated.

## Data Availability

All relevant data are within the manuscript and its Supporting Information files.

## Acknowledgements

None.

